# Introduction of a Penicillin Allergy De-Labelling Program with Direct Oral Challenge is Associated with Increased Downstream Utilization of Beta-Lactam Antimicrobials: A Multicenter Parallel Cohort Study with Crossover Evaluation

**DOI:** 10.1101/2023.05.29.23290698

**Authors:** Adhora Mir, Derek Lanoue, Veronica Zanichelli, Carl van Walraven, Timothy Olynych, Caroline Nott, Derek MacFadden

## Abstract

**Background:** Penicillin allergy labels are common and are often inaccurate. These labels can lead to unnecessary use of second-line non-beta-lactam antibiotics, and worse clinical outcomes.

**Objectives:** We measured the impact of the introducing of a standardized proactive penicillin allergy de-labelling program with oral amoxicillin challenge on subsequent antibiotic use.

**Methods:** We performed a retrospective comparison of parallel cohorts from two separate tertiary care hospital campuses across two penicillin de-labelling intervention periods. Outcomes included data including penicillin allergy label and antibiotic use, were collected for the index admission and the subsequent 6-month period. Descriptive statistics as well as multivariate regression analyses were performed.

**Results:** A total of 368 patients with penicillin allergy label were included across two campuses and study periods. 24 (13.8%) patients in the intervention group sustained penicillin allergy label at 30 days from admission vs. 3 (1.5%) in the non-intervention group (p < 0.001). In the 6-months following admission, beta-lactams were prescribed more frequently in the intervention groups vs. the non-intervention groups for all patients (28 [16.1%] vs 15 [7.7%], p= 0.04) and for only those patients who received antibiotics (28/46 [60.9%] vs. 15/40 [37.5%], p=0.097). In a multivariate analysis, the intervention was found to be associated with an increased odds of beta-lactam prescribing in all patients (OR 2.49, 95% CU 1.29-5.02) and in those prescribed at least one antibiotic (OR 2.44, 95% CI 1.00-6.15). There were no differences in overall antibiotic prescribing by intervention and non-intervention group during admission (113 [64.9%] vs. 112 [57.7%]) or within 6-months (46 [26.4%] vs. 40 [20.6%]). No drug related adverse events were reported.

**Conclusions:** Proactive penicillin allergy de-labelling for inpatients was associated with a reduced number of penicillin allergy labels and increased utilization of beta-lactam vs. other antibiotics in the subsequent 6-months.

**Capsule Summary:** A proactive systematic approach to antibiotic allergy de-labelling for inpatients with penicillin allergy label results in an increased number of patients de-labelled at hospital discharge and increased beta-lactam use in the subsequent 6 months.

## INTRODUCTION

Approximately 10% of inpatients report a penicillin allergy, but studies have shown that over 90% of these patients will tolerate a penicillin-based antibiotic [2].

Moreover, 43% of patients are identified as having “low risk” histories and can be delabeled safely through direct penicillin oral challenge [1]. Avoidance of penicillin-based antibiotics due to penicillin allergy labels leads to unnecessary use of second line agents, glycopeptides, fluoroquinolones, lincosamides, or aminoglycosides, that can be less effective, have a greater risk of side effects, and be costlier than betalactams [3-4]. Therefore, penicillin de-labelling programs are an important antimicrobial stewardship tool [1]. However, their uptake has been suboptimal because of the historic need for penicillin skin-testing which are labour-intensive, costly, and require specialist input [5].

Recent studies have demonstrated the safety and efficacy of oral challenge of penicillin-based antibiotics for inpatients with remote, low-risk, cutaneous-only reactions [6-8]. These low-risk patients have histories of reactions that are strictly cutaneous, involve vague symptoms, or consist of unknown manifestations. In a retrospective review of military recruits undergoing direct oral amoxicillin challenge, 0/328 (0%) and 5/328 (1.5%) experienced an anaphylactic or any reaction, respectively [9]. Ramsey *et al*. demonstrated the efficacy of oral challenge of penicillin-based antibiotics in the inpatient and outpatient setting in patients with lowrisk, cutaneous-only reactions occurring more than 10 years ago [7,10]. Confino-Cohen and colleagues challenged a total of 617 patients with a history of non-immediate reactions regardless of skin test results and only 9 patients (1.5%) experienced an immediate reaction, all of which were mild [11]. Mill *et al*. demonstrated an exceptional safety profile of direct challenges in the pediatric population with a history of cutaneous reactions [12]. While direct oral challenge appears safe, real-world data are needed to support the feasibility of implementation in the inpatient setting and the downstream impacts of de-labelling programs, including subsequent utilization of beta-lactam antibiotics.

This study measured the expected benefit of introducing a standardized de-labelling program at two large campuses of a tertiary care academic health center.

## METHODS

### Study Design

A penicillin allergy delabeling program was previously implemented at a large tertiary care center (The Ottawa Hospital) in Ottawa Canada as part of a quality improvement initiative during two separate time periods at two campuses. We performed a retrospective, parallel cohort with crossover study, to measure its effect on penicillin allergy de-labelling and antibiotic prescribing in the subsequent 6-month period.

Data for this study was collected retrospectively from across two time periods defined as Period 1 (April 15th to April 30th 2021), and Period 2 (February 15th to March 8th 2022). During Period 1, the intervention occurred at Campus B but not Campus A, and during Period 2 the intervention occurred at Campus A but not Campus B (Figure 1), creating a natural parallel cohort with crossover. Ethics approval from the Ottawa Hospital was obtained for this retrospective study. The prior described quality improvement initiative had an REB exemption at the Ottawa Hospital. STROBE guidelines were followed during the development, analysis, and reporting of this observational study [13].

**Figure 1:**
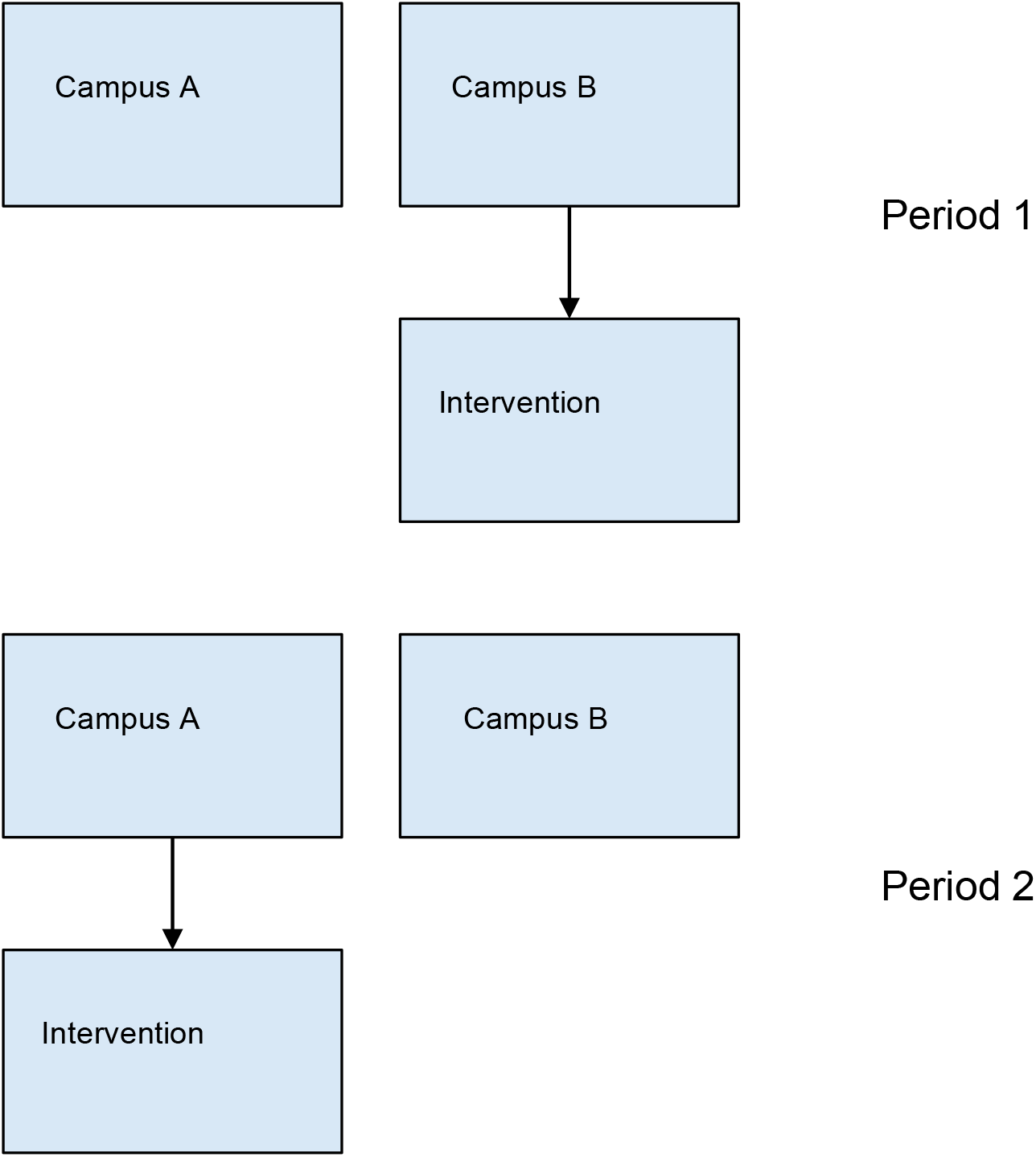
Schematic representation of the de-labelling intervention.

### The Penicillin De-labelling Program with Oral Amoxicillin Challenge

A penicillin allergy de-labelling with oral amoxicillin challenge quality improvement initiative was previously implemented during two time periods at two separate campuses as previously noted. We have described this program in detail in the supplemental methods section, as well as in brief herein. The program consisted of daily systematic screening of adult hospital inpatients admitted to medical or surgical services to identify patients with a penicillin allergy label documented in the electronic medical records (EPIC). Patients meeting eligibility criteria were further evaluated to assess their risk of true penicillin allergy, and a management algorithm was applied. Low-risk patients were identified and offered amoxicillin oral challenge (250 mg oral dose x 1) if they met eligibility requirements for the procedure and with approval from their responsible team. Moderate and high risk patients were referred to an allergy and immunologist for further evaluation as an outpatient. Some patients, with a family history of penicillin allergy but no personal history of a penicillin allergy or drug intolerance had their penicillin label directly removed from their chart. If patients could not be classified, they were reviewed at weekly meetings with investigators and board-certified Clinical Immunologist and Allergist to determine appropriate allergy testing group placement.

### Population

We included inpatients ≥ 18 years admitted to a non-psychiatric hospital bed for >24h to a medical or surgical service who had a reported penicillin allergy label listed in EPIC electronic medical record system were identified and screened by one of the investigators. During participant screening, each person’s record was reviewed to identify the presence of any exclusion criteria including: a) pregnancy; b) respiratory or hemodynamic instability (SBP<100, HR>120, need for vasopressors, requiring > 4L/min oxygen); c) documented history of active suicidal ideation, dementia or current delirium; d) active COVID-19 infection. The latter criterion was included for infection control reasons. Those with same day surgical admission were excluded for the purposes of this study. Patients without exclusion criteria were approached by the assessor after agreement from the patient’s most responsible physician (MRP).

### Outcomes

The two primary outcomes were: (1) removal of penicillin allergy label at 30 days from admission; and (2) receipt of beta-lactam antibiotics as an in-patient at our hospital within 6-months of admission, respectively. Secondary outcomes included: (1) the use of any antibiotic on initial admission and within 6-months as an in-patient at our hospital, (2) the use of beta-lactam antibiotics on initial admission and within 6-months as an in-patient at our hospital for all patients, and amongst only those patients who received an antibiotic; (3) the presence of penicillin allergy label in the EMR at 48 hour and 6-month time points from admission; (4) the use of non-beta-lactam antibiotics on initial admission and within 6-months as an in-patient at our hospital for all patients, and amongst only those patients who received an antibiotic; and (5) the prevalence of *C*.*difficile* by 3-months.

### Covariates

Covariates included: (1) demographics (Age and Sex); (2) admitting service (Medical or Surgical); (3) comorbidities via the Charlson comorbidity score [14]; (4) number of non-penicillin allergy labels; and (5) use of systemic antibiotics at our hospital within 6-months prior to admission; and (6) mean length of stay.

### Statistical Analysis

Descriptive statistics were presented as counts and continuous variables, and summarized as proportions and means/medians. We compared count variables using chi-square testing, and continuous variables via t-test. Descriptive statistics were stratified by relevant covariates. We used multivariable logistic regression modeling to calculate effect estimates of intervention on the primary outcome and selected secondary outcomes after adjusting for the aforementioned covariates including campus. Statistical analysis was performed using RStudio version 2022.07.2+576 software.

## RESULTS

A total of 368 patients were included across the two campuses and study periods. For both campuses, mean age ranged from 60-61 years, and the majority of patients were female (63%-72%). Most patients were admitted under medical services (51-55%) and had more than 1 allergy label (Table 1). Sex, age, admitting service, antibiotic prescribing in the prior 6 months, and number of non-penicillin allergy labels and Chalrson morbidity index score were not significantly different (Table 1) between intervention and non-intervention groups.

**Table 1.** Baseline characteristics by hospital and intervention periods.

|  | Intervention |  | Non-Intervention |  | Total |  |
| --- | --- | --- | --- | --- | --- | --- |
|  | Campus A (N=74) | Campus B (N=100) | Campus A (N= 107) | Campus B (N= 87) | Campus A (N=181) | Campus B (N= 187) |
| **Sex** |  |  |  |  |  |  |
| Female | 50 (67.6%) | 75 (75.0%) | 64 (59.8%) | 60 (69.0%) | 114 (63.0%) | 135 (72.2%) |
| Male | 24 (32.4%) | 25 (25.0%) | 43 (40.2%) | 27 (31.0%) | 67 (37.0%) | 52 (27.8%) |
| **Age** |  |  |  |  |  |  |
| Mean (SD) | 57.4 (21.6) | 61.5 (20.2) | 62.3 (17.6) | 59.8 (18.7) | 60.3 (19.4) | 60.7 (19.5) |
| Median [Min, Max] | 59.0 [20.0, 99.0] | 64.0 [21.0, 99.0] | 64.0 [20.0, 95.0] | 64.0 [21.0, 96.0] | 62.0 [20.0, 99.00] | 64.0 [21.0, 99.0] |
| **Charlson score** |  |  |  |  |  |  |
| Mean (SD) | 1.04 (1.50) | 1.63 (2.59) | 1.32 (1.81) | 1.67 (2.55) | 1.20 (1.69) | 1.65 (2.56) |
| **Admitting service** |  |  |  |  |  |  |
| Medical | 38 (51.4%) | 49 (49.0%) | 54 (50.5%) | 53 (60.9%) | 92 (50.8%) | 102 (54.5%) |
| Surgical | 36 (48.6%) | 51 (51.0%) | 53 (49.5%) | 34 (39.1%) | 89 (49.2%) | 85 (45.5%) |
| **Number of other allergies** |  |  |  |  |  |  |
| Mean (SD) | 1.18 (1.58) | 1.45 (1.94) | 1.51 (2.03) | 1.68 (2.45) | 1.38 (1.86) | 1.56 (2.19) |
| **Antibiotics used 6 months prior to admission** |  |  |  |  |  |  |
| Antibiotics Used | 14 (18.9%) | 12 (12.0%) | 21 (19.6%) | 19 (21.8%) | 35 (19.3%) | 31 (16.6%) |
| No Antibiotics Used | 60 (81.1%) | 88 (88.0%) | 86 (80.4%) | 68 (78.2%) | 146 (80.7%) | 156 (83.4%) |

The primary outcome was the number of patients whose penicillin allergy was delabeled 30 days from admission, and significantly more patients were de-labelled in the intervention group (24 [13.8%]) than the non-intervention group (3 [1.5%]) ((p<0.001) (Table 2). Of the 24 patients de-labelled in the intervention arm, 19 had received a direct oral challenge. No significant drug reactions were reported.

**Table 2.** Unadjusted primary outcomes by intervention periods

|  | Intervention (N=174) | Non-Intervention (N=194) | Total (N=368) | P-Value | OR (95% CI) |
| --- | --- | --- | --- | --- | --- |
| **Patients delabeled 30 days from admission** | 24 (13.8%) | 3 (1.5%) | 27 (7.3%) | <0.001 | 10.19 (3.01-34.47) |
| **Beta-lactam use 6 months from admission\*** | 28 (60.9%) | 15 (37.5%) | 43 (50.0%) | 0.0966 | 2.59 (1.08-6.20) |

\*Among those patients who received any antibiotic.

During the index admission, 113 (64.9%) and 112 (57.7%) patients received an antibiotic in the intervention and non-intervention groups respectively. Among these patients, 63 (55.8%) and 64 (57.1%), received beta-lactams in the intervention and non-intervention group, respectively (p=0.98) (Table 3). As well, there were no differences in the total number of patients receiving any antibiotic within 6-months between intervention (46 [26.4%]) and non-intervention (40 [20.6%]) groups. Of patients who received an antibiotic prescription during the 6-months following admission, beta-lactams were prescribed more frequently in the intervention groups 28 (60.9%) compared with the non-intervention groups 15 (37.5%) (Table 3). After adjusting for potential confounding factors with multivariable logistic regression, the intervention was found to be significantly associated with increased beta-lactam use in the 6-months following index admission in those that used antibiotics (OR 2.44, 95% CI 1.00-6.15, p=0.05) (Table 4). When we expanded the analysis to all patients, the effect was more pronounced (OR 2.49, 95% CI 1.29-5.02, p= 0.008).

**Table 3.** Unadjusted secondary outcomes by intervention periods

|  | Intervention (N=174) | Non-Intervention (N=194) | Total (N=368) | P-Value | OR (95% CI) |
| --- | --- | --- | --- | --- | --- |
| **Patients delabeled 48 hours from admission** | 23 (13.2%) | 2 (1.0%) | 25 (6.8%) | <0.001 | 14.63 (3.39-62.99) |
| **Patient delabeled 6 months from admission** | 26 (14.9%) | 3 (1.5%) | 29 (7.9%) | <0.001 | 11.18 (3.32-37.67) |
| **Beta-lactam use during admission\*** | 63 (55.75%) | 64 (57.1%) | 127 (56.4%) | 0.98 | 1.49 (0.86-2.56) |
| **Non Beta-lactam use during admission\*** | 76 (67.3%) | 65 (58.0%) | 141 (62.7%) | 0.36 | 0.95 (0.56-1.60) |
| **Non Beta-lactam use 6 months from admission\*** | 24 (52.2%) | 29 (72.5%) | 53 (61.6%) | 0.15 | 0.41 (0.17-1.02) |
| **Beta-lactam use during admission** | 63 (36.2%) | 64(34.0%) | 129(35.1%) | 0.91 | 1.10 (0.72-1.69) |
| **Beta-lactam use 6 months from admission** | 28 (16.1%) | 15 (7.7%) | 43 (11.7%) | 0.04 | 2.29 (1.18-4.45) |
| **Non Beta-lactam use during admission** | 76 (43.7%) | 65 (34.5%) | 143 (38.9%) | 0.2 | 1.47 (0.96-2.24) |
| **Non Beta-lactam use 6 months from admission** | 24 (14.4%) | 29 (14.9%) | 54 (14.7%) | 0.99 | 0.95 (0.53-1.70) |
| **C. difficile infection 3 months from admission** | 2 (1.1%) | 4 (2.1%) | 6 (1.6%) | 0.79 | 0.55 (0.10-3.05) |
| **Mean length of stay (SD)** | 6.93 (8.49) | 7.74 (10.8) | 7.35 (9.77) | 0.42 | NA |

\*Among those patients who received any antibiotic.

**Table 4.** Multivariable adjusted odds ratios by predictor and outcome

|  | Patients delabeled 30 days from admission | Beta-lactam use 6 months from admission (antibiotic users) | Beta-lactam use 6 months from admission (all comers) |
| --- | --- | --- | --- |
| Intervention Status | 12.34 (4.12-53.35) | 2.44 (1.00-6.15) | 2.49 (1.29, 5.02) |
| Campus | 4.25 (1.76-11.47) | 0.89 (0.34-2.30) | 1.68 (0.87-3.17) |
| Sex | 0.99 (0.39-2.68) | 1.26 (0.45-3.27) | 0.85 (0.42-1.75) |
| Age | 1.00 (0.98-1.03) | 1.01 (0.98-1.04) | 1.00 (0.98-1.02) |
| Charlson | 1.03 (0.82-1.37) | 1.07 (0.86-1.34) | 0.99 (0.86-1.18) |

Non-beta lactam use during admission among those who used antibiotics did not significantly differ between intervention and non-intervention groups, occuring in 76 (67.3%) patients in the intervention group and 65 (58%) in the non-intervention group (Table 3). There was a trend to more patients in the non-intervention group (29 [72.5%]) having received non-beta lactam antibiotics in the 6 months following admission than the intervention group (24 [52.2 %]), although this was not significant (p=0.15).

There was no significant difference in the incidence of *C*.*difficile* infection at 3 months 1.2% (n=2) and 2.1% (n=4), amongst the intervention and non-intervention groups respectively.

Mean length of stay in the intervention group (6.93 days) was reduced compared to mean length of stay in the non-intervention group (7.74 days), however this difference was not significant when t-test was applied (p=0.42).

## DISCUSSION

In this study we found that implementation of a systematic penicillin allergy screening and direct oral challenge program was associated with both increased removal of penicillin allergy labels at 30 days from admission, and a greater utilization of betalactams when patients were prescribed antibiotics within the 6-months after the intervention period. Furthermore, there were no adverse events of direct oral challenge reported from the original direct oral challenge intervention. Introduction of de-labelling programs to the inpatient setting appears safe, effective, and can help patients preferentially receive beta-lactam antibiotics, typically the first line class of antibiotics.

Our study complements other studies in the literature, showing the effectiveness, safety, and effect on antibiotic prescribing of an inpatient penicillin allergy assessment program [6-12]. Recent studies by Ramsey et al. [7] and Chua et al. [8] support the safety of this direct oral challenge approach, and it is the standard of care for assessment of low risk penicillin allergies outlined in the current North American drug allergy guidelines [2].

Few studies have evaluated the downstream consequences of de-labelling on antibiotic prescribing [8, 15, 16], and typically do so by evaluating only those outcomes in individual patients receiving the specific intervention. In this paper, we demonstrate a significant impact at the level of all patients with penicillin allergy label who were located at a hospital receiving the de-labelling intervention. These provide compelling evidence for broader adoption of these approaches and potential implementation as part of antimicrobial stewardship program elements [17].

Commensurate with the aim of improving beta-lactam usage, we did see a trend towards reduced proportional non-beta-lactam use in intervention group compared to the non-intervention group in the 6-month after admission, which fits with the expected replacement of non-beta-lactam antibiotics by beta-lactam antibiotics on those who were delabelled. While there was no significant effect of intervention on incidence of *C. difficile* in the 3 months following admission, the outcome was rare.

Initial admission length of stay was not markedly impacted by intervention group, but may have a delayed impact on length of stay in subsequent visits, and warrants further exploration.

Our study has several limitations. First, is that the original de-labelling intervention excluded a large number of inpatients prior to assessment. Many of the potentially eligible patients were quite unwell with 56% of patients being excluded from assessment based on pre-defined exclusion criteria. The primary reason for exclusion were cognitive issues that would prevent accurate assessment such as delirium, dementia or active suicidal ideation. In spite of these exclusions, we still found a marked beneficial impact of the intervention at the population level. As sample size was limited in our study, larger studies at other centers will be needed to definitively measure the effect of delabelling on outcomes. A second limitation to the study was that it was retrospective in nature and without a placebo control group. A trial in which eligible patients were randomized to delabelling vs standard care would have let us directly measure the influence of oral challenge on outcomes. Our non-randomized study design is unable to determine whether outcomes differing between the intervention group and observational controls was due to the challenge vs. the entire intervention.

While this does pose some limitations to inference, the study did benefit by the ability to compare two parallel groups that changed intervention periods, which may yield a natural balancing of confounders beyond those measured and adjusted for in the analyses.

In summary, our study supports the adoption of inpatient programs for penicillin allergy de-labelling with direct oral challenge to help reduce the number of inappropriate penicillin allergy labels in patients and enable improved future utilization of first line beta-lactam antibiotics. Future studies are needed to answer if these interventions result in improvements across other patient outcomes including length of stay, antibiotic toxicities, and even infection related mortality.

## SUPPLEMENTAL MATERIALS

### Procedures

Over two distinct 2 week periods, the physician investigator used a daily report generator on the *Epic* Electronic Medical Record system to identify all patients admitted to any medicine or surgery service at the hospital in the last 24 hours who were labeled with penicillin allergy. Patients were approached by the investigator after agreement by the patient’s most responsible physician (MRP). Patients with documented history of pregnancy, dementia, active suicidal ideation or current delirium were be assessed separately and shared decision between primary treatment team and patient or SDM to participate will be established before proceeding. The investigator then used the Penicillin Allergy De-Labeling Algorithm approved by the Ottawa Hospital Antimicrobial Subcommittee to identify patients having a low-risk history of penicillin reaction (Version date 6 April 2021). The inclusion criteria are outlined in figure 2.

**Supplemental Figure 1:**
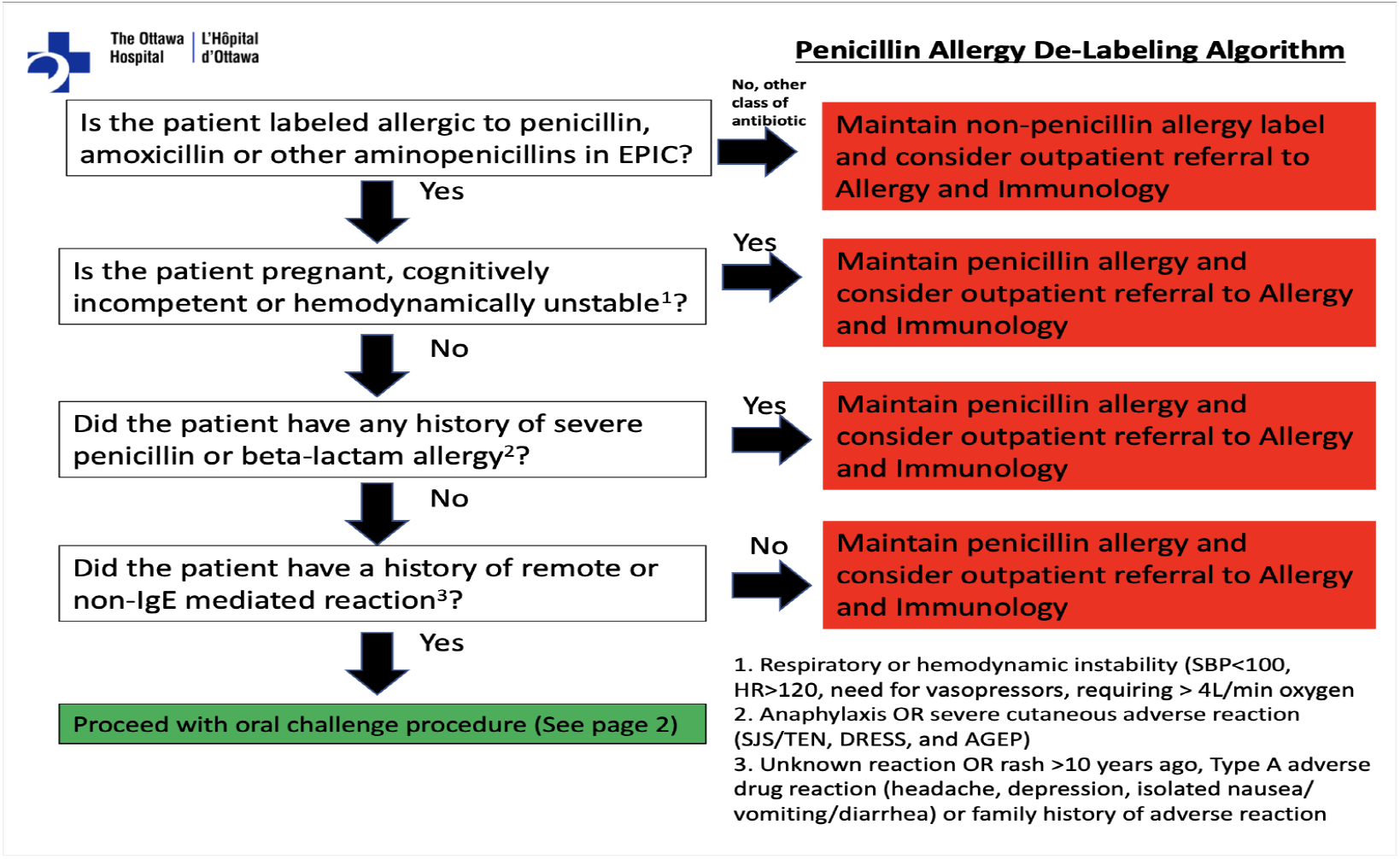
Risk stratification algorithm

Those with any history suggestive of a severe, non Ig-E mediated reaction to penicillin (blistering, mucous membrane involvement, fever, joint pain/swelling) were told to continue to avoid penicillin’s. Patients with a high risk history of penicillin reaction (anaphylaxis, oropharyngeal angioedema, wheezing, hemodynamic alteration, any reaction within 1 year) were referred to Allergy and Immunology for further testing consideration. Similarly, patients with a moderate risk history of penicillin reaction (itching, hives, non-specific rash 1-5 years ago, time frame of reaction not clearly beyond this interval, any reported angioedema, recall of need for urgent medical attention) were referred to Allergy and Immunology.

Patients with a low risk history of penicillin reaction (cutaneous reaction >10 years ago, rash or doesn’t remember, no urgent medical attention needed, or patient does not know details of reaction occurring >5 years ago) underwent informed consent for oral challenge to a penicillin.

As well, patients with family history of penicillin allergy but no personal history of a penicillin allergy or drug intolerance had their penicillin label directly removed from their chart. If patients could not be classified, using the above algorithm, they were reviewed at weekly meetings with investigator and board-certified Clinical Immunologist and Allergist to determine appropriate allergy testing group placement.

All patients electing to proceed with de-labelling provided informed consent. The patients were then administered an oral challenge consisting of 250mg PO amoxicillin followed by 60 minutes of monitoring by direct supervision by the investigator on the hospital ward. Patients will be monitored for adverse reactions including anaphylaxis. The occurrence of any of these symptoms or signs will have classified the person as having had an adverse drug event. Patients without any of these findings were classified as ‘de-labelled patients’ and had their allergy status corrected on the electronic medical record system. In addition, a note was sent to their primary care provider and pharmacy regarding their corrected penicillin allergy status.

**Supplemental Figure 2:**
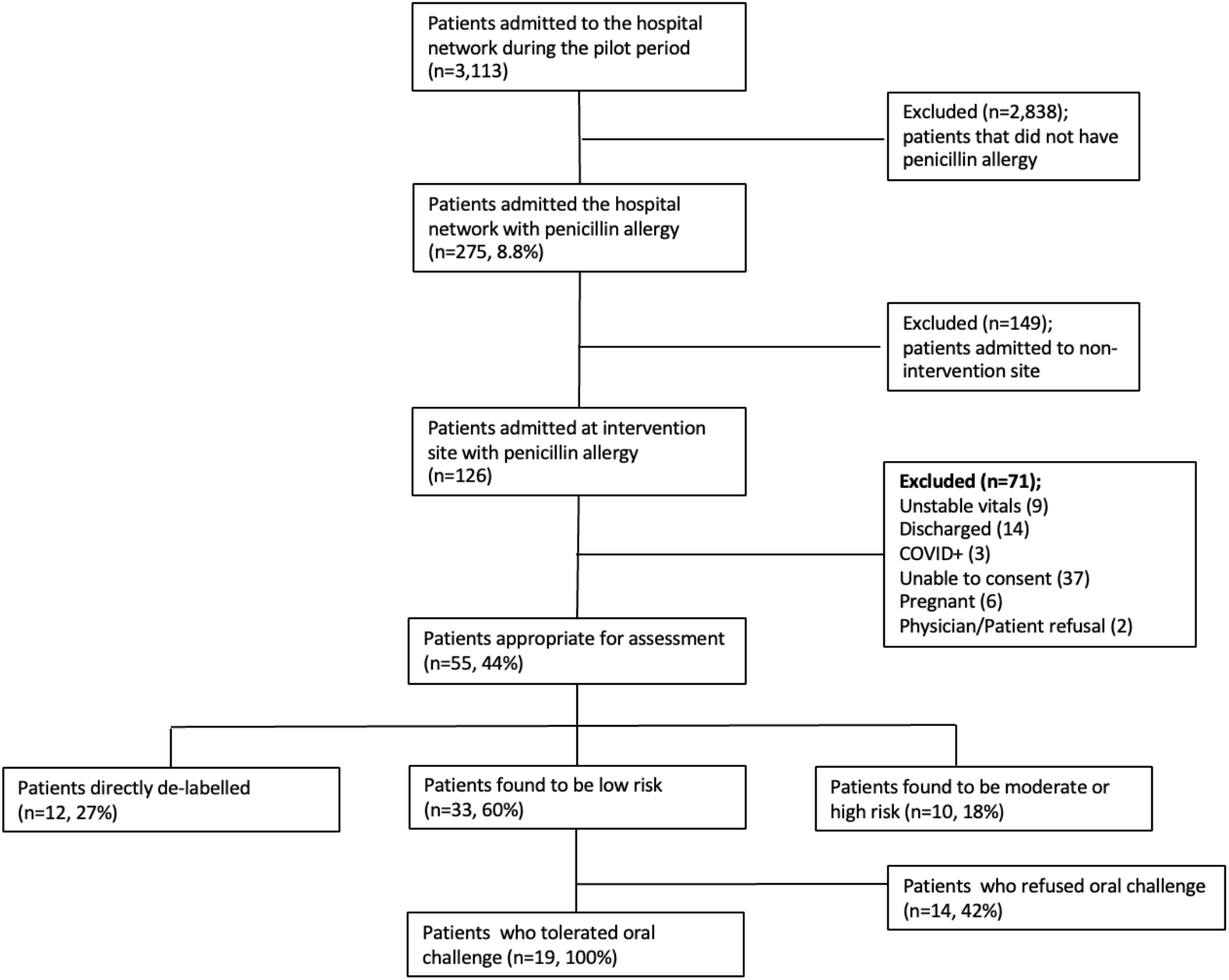
Patient Assessment Data

### Limitations

When comparing the data pulled from the electronic medical record and investigator records, 7 patients that were de-labeled were not captured from the data pull. These patients were explained by delays in the updating of allergy labels on the electronic medical record based on nursing acknowledgement of label removal. This suggests the true effect of the study may be greater than above analysis.

## Data Availability

All data produced in the present study are available upon reasonable request to the authors

## Abbreviations

EMR: (electronic medical record)
ASP: (Antimicrobial Stewardship Program)
TOH: (The Ottawa Hospital)
SBP: (Systolic blood pressure)
HR: (heart rate)
DMARD: (disease modifying antirheumatic drugs)

## AKNOWLEDGEMENTS

This project was supported by an Innovation grant awarded by The Ottawa Hospital Academic Medical Organization (TOHAMO).

## Notes

**Clinical Implications.** These results support the adoption of a hospital specific pro-active program to aid penicillin allergy de-labelling including direct oral challenge and support beta-lactam antibiotic use where indicated.

### Competing Interest Statement

The authors have declared no competing interest.

### Author Declarations

Ottawa Health Science Network Research Ethics Board (OHSN-REB) waived ethical approval for this work.

